# Effectiveness of endurance training rehabilitation after hospitalisation in intensive care for COVID-19-related acute respiratory distress syndrome on dyspnoea (RECOVER): a randomised controlled, open-label multicentre trial

**DOI:** 10.1101/2022.08.29.22279327

**Authors:** Christophe Romanet, Johan Wormser, Audrey Fels, Pauline Lucas, Camille Prudat, Emmanuelle Sacco, Cédric Bruel, Gaëtan Plantefève, Frédéric Pene, Gilles Chatellier, François Philippart

## Abstract

**Background:** COVID-19-related Acute Respiratory Distress Syndrome (CARDS) is the severe evolution of the Sars-Cov-2 infection leading to an intensive care unit (ICU) stay. Its onset is associated with “long-covid” including persisting respiratory disorders up to one year. Rehabilitation is suggested by most guidelines in the treatment of “long-covid”. As no randomised controlled trial did support its use in “long-covid” we aimed to evaluate the effects of endurance training rehabilitation (ETR) on dyspnoea in “long-covid” following CARDS.

**Methods:** In this multicentre, two-arm, parallel, open, assessor-blinded, randomised, controlled trial performed in three French ICU, we enrolled adults previously admitted for CARDS, discharged for at least three months and presenting an mMRC dyspnea scale score > 1. Eligible patients were randomly allocated (1:1) to receive either ETR or standard physiotherapy (SP), both for three months. Outcomes assessors were masked to treatment assignment. Primary outcome was dyspnoea’s evolution, measured by Multidimensional Dyspnea Profile (MDP) at inclusion and after 90 days.

**Results:** Between August 7, 2020 and January 26, 2022, 871 COVID-19 patients were screened, of whom 60 were randomly assigned to ETR (n=27) or SP (n=33). Mean MDP score after treatment was significantly lower in the ETR group than in the SP group (26.15 [SD 15.48] vs. 44.76 [SD 19.25]; mean difference -18.61 [95% CI -27.78 to -9.44]; p<0.0001).

**Conclusion:** CARDS patients suffering from breathlessness three months after discharge improved their dyspnoea significantly more when treated with ETR for three months rather than with SP.

## Introduction

The COVID-19 pandemic has challenged the worldwide healthcare system by a surge of patients admitted to hospitals for acute respiratory failure (ARF). Although for the majority of patients the SARS-CoV-2 infection ended naturally, 10% eventually developed an hyperinflammatory state leading to severe hypoxemia that would require mechanical ventilation (MV) due to the development of a COVID-19-related Acute Respiratory Distress Syndrome (CARDS) associated with an overall 90-day mortality of about one third of patients [1, 2].

While CARDS has a broad similarity with ARDS [3], several clinical and physiopathological patterns raise questions about the relevance of usual therapeutic guidelines [4]. An “Happy hypoxemia”, corresponding to a profound decrease in oxygen saturation observed in remarkably non-dyspnoeic patients has been reported repeatedly in COVID-19 patients [5, 6]. In addition, while ARDS is associated with a loss of lung compliance, initial CARDS often combines severe hypoxaemia with near-normal lung compliance. This unprecedented observation in ARDS led to the development of two quite contrasting phenotypes of CARDS (Type H, Type L), only sharing their severe hypoxemia [7]. Finally, the use of methylprednisolone, of potential interest during classical ARDS and initially suggested as an interesting modulator of the lungs’ hyperinflammatory response may have been outperformed by dexamethasone, which has been shown to have superior effects in CARDS [8, 9].

Following ARDS, close to 50% of patients develop neuromuscular dysfunction labelled “ICU-acquired weakness” (ICU-AW) affecting symmetrically limbs (muscle weakness) and the respiratory muscles (dyspnoea) [10, 11]. Functional disability, muscle weakness and dyspnoea persist for years following ICU and hospital discharge, dramatically affecting quality of life [12, 13]. Similar clinical patterns seem to be observed during post-CARDS “long-covid” evolution [14–17].

International recommendations did stress the importance of physiotherapy and rehabilitation in “long-covid” to treat the remaining symptoms, especially dyspnoea and functional disability [18, 19]. However, although physiotherapy during ICU stay and after discharge is well described and seems effective despite a highly heterogeneous literature, rehabilitation after ICU discharge has on the other hand hardly been studied; both seem to be promising curative treatments, but high quality and power randomised controlled trials are still lacking [20]. Furthermore, due to the significantly different and still not fully understood course of CARDS, patients may be exposed to harsher physical, respiratory and psychological complications compared to ARDS [21, 22]. However the specific physio-pathological mechanisms may lead to a peculiar response to rehabilitation [21].

We therefore sought to assess the effects of endurance training rehabilitation (ETR) on dyspnoea and quality of life compared to standard physiotherapy (SP) care amongst post-CARDS “long-covid” patients.

## Methods

### Study design and participants

This investigator-initiated, multicentre, randomised–controlled, two-arm, parallel, open-label, assessor-blinded trial took place in three intensive care units in Paris, France.

Adults with CARDS were eligible for recruitment if they had received mechanical ventilation for at least 48 hours; had been discharged from ICU for at least three months; reported dyspnoea with a modified Medical Research Council (mMRC) dyspnoea scale above one at the time of inclusion. Patients were excluded if they were unable to take part in rehabilitation sessions (severe neurological diseases, osteoarticular pathology) or in case of excessive geographical distance (>5km) to the rehabilitation practice. Full inclusion and exclusion criteria are provided in the appendix.

Participants were screened and informed via phone call by the research physiotherapist (CR). According to the French law (Loi Jardé), the oral consent of each participant was obtained and recorded in the patient’s electronic medical record. Inclusion was then made the following week by the doctor in charge of outcomes’ assessment (PL). The trial was conducted in accordance with the principles stated in the Declaration of Helsinki and Good Clinical Practice guidelines. The protocol has been approved by an Ethics Committee (Comité de Protection des Personnes Sud-Méditerranée III, final approval 29 june 2020, RCB 2020-A01686-33) and was registered on Clinicaltrials.gov (NCT04569266) on September 29, 2020.

### Randomisation

Screened patients meeting the eligibility criteria were selected consecutively.

Eligible patients were randomly allocated (1:1) to receive either ETR or SP, both for a duration of three months (two sessions per week). The trial statistician generated a permuted-block randomisation sequence using variably sized blocks of two or four with stratification according to centre and sequentially assigned each participant to his/her intervention.

### Procedures

Patients in the intervention group received a prescription for ETR, at the rate of two one-hour sessions per week for 10 weeks, according to guidelines [23]. Patients in the control group received a prescription for SP, at the rate of two sessions per week for 10 weeks. The investigator who completed outcomes assessments at inclusion and at trial’s end (PL) was blinded from randomisation and group allocations. Physiotherapists giving ETR or SP were external practitioners and did not take any part in study design, patient selection or results analysis. All physiotherapists were independent of the three recruiting centres.

In the ETR group, continuous endurance training was planned in accordance with guidelines [23]. Details of this intervention are given in the appendix.

Patients allocated to the SP group benefited from the same number of sessions. As no guidelines exist about SP, no specific recommendations were given to the physiotherapists. Of note, at the end of study period, an ETR prescription was given to the SP group, considering the current guidelines in COVID-19 patients [24].

### Outcomes

The primary outcome was the evolution of dyspnoea, assessed by the difference in the Multidimensional Dyspnea Profile (MDP) score between baseline and at 90 days from inclusion (end of intervention period).

Secondary outcomes were the evaluation of dyspnoea by the Modified Medical Research Council (mMRC) scale and measurement of quality of life evolution by the 12-item Short-Form Survey (SF-12) at 90-days from inclusion.

### Sample size

At the date of protocol implementation, there was only little data available on the Minimal Clinically Important Difference, especially in this peculiar population. We therefore chose a MDP MCID of 12 points obtained by anticipating a change of more than one point per item, with a standard deviation of 30 points. Using these values, a Student t-test, a two-sided alpha of 0.05, and 90% power, we estimated that 100 persons would need to be enrolled in each group.

### Statistical analysis

According to their distribution, continuous variables are presented as mean (standard deviation) or median (interquartile range), and categorical data as numbers (percentage).

Concerning the three-month MDP (primary outcome), between-groups differences were tested using an ANCOVA model adjusted for baseline values. Secondary outcomes were analysed in the same way. All estimates are presented with their 95% confidence interval.

We performed all analyses with the R 4.2.1 software (The R Project for Statistical Computing, https://www.r-project.org), according to the intention-to-treat principle, considering a two-sided type I error with an alpha of 0.05.

## Results

Between July 22, 2020 and January 26, 2022, 871 patients were assessed for eligibility of whom 60 underwent randomisation: 27 patients in the ETR group and 33 in the SP group. Most patients were recruited in the Groupe Hospitalier Paris Saint-Joseph (n=32, 55.17%). 811 patients (93%) did not fulfil inclusion criteria at initial screening. All included patients followed the entire protocol training until its end. None withdrew consent after randomisation and none dropped out of the training program. Four patients assigned to the ETR group received the SP program, while three patients assigned to the SP program received the ETR program.

All patients completed the last follow-up assessment. The last follow-up visit took place on March 25, 2022. The study was closed on March 28, 2022 due to the phasing out of ICU admissions for ARDS in COVID-19 patients. The mean time between ICU discharge and inclusion were 173 (95% CI 147.36 to 198.64) and 174 (95% CI 144.42 to 204.92) days in the ETR and SP groups, respectively. Full details on reasons for patients’ exclusion and their follow-up are shown on Figure 1.

**Figure 1.**
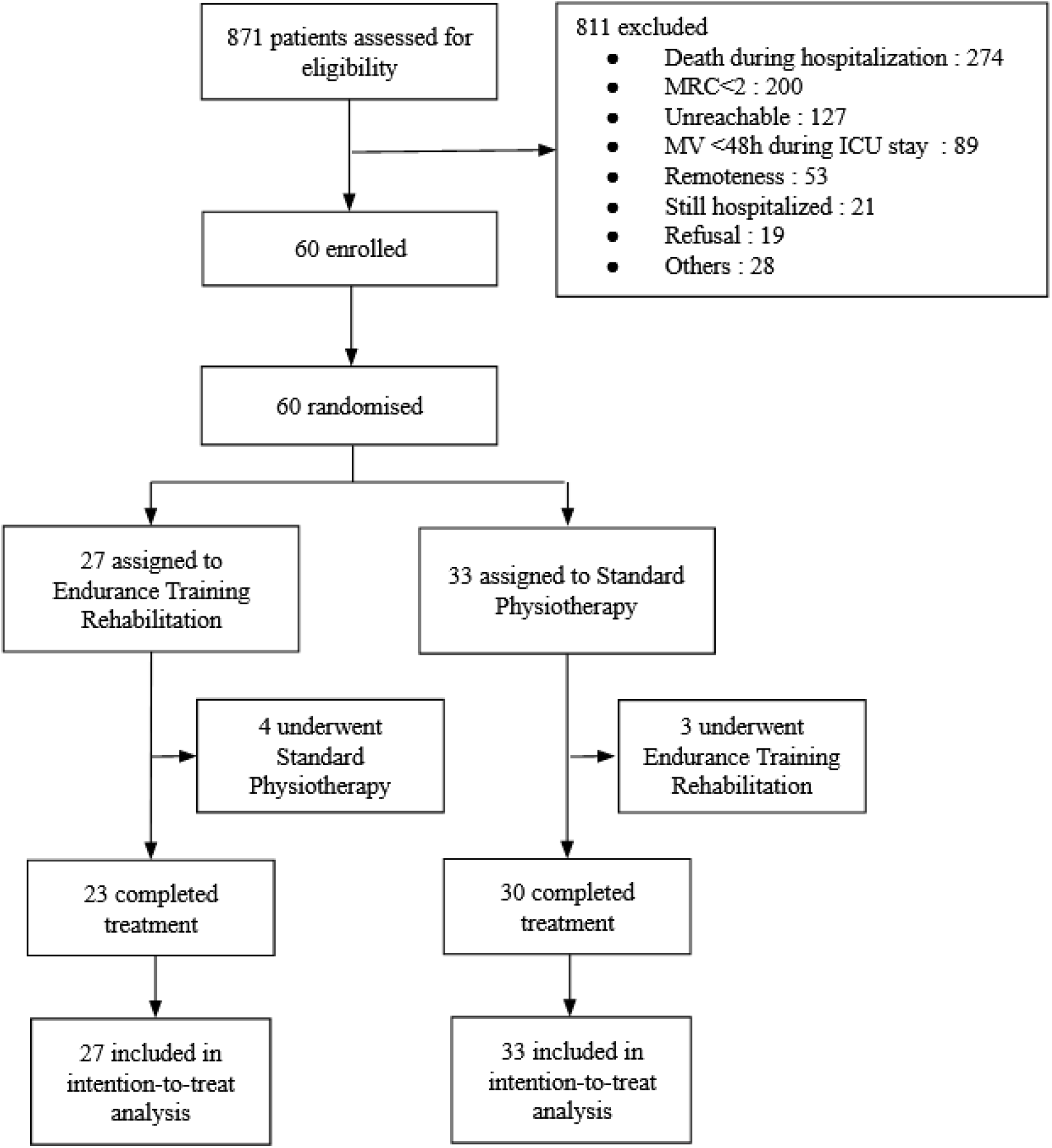
Trial profile: eligibility assessment, randomization, follow-up and analysis.

Demographic and baseline characteristics were comparable between groups. The mean age of patients was 58.15 years (SD 12.05), 37 were men (61.67%) and 23 were women (38.33%). The main comorbidity was diabetes (n=22, 36.67 %). Full details about baseline characteristics of patients are presented in table 1.

**Table 1:** Baseline characteristics of patients. Data are mean (SD), n (%) or median (IQR Q1-Q3) Abbreviations: ETR: endurance training rehabilitation; SP: standard physiotherapy

|  | ETR (n=27) | SP (n=33) |
| --- | --- | --- |
| **Demographics** |  |  |
| Age, years | 56.67 (14.28) | 59.36 (9.94) |
| Women | 11 (40.74%) | 12 (36.36%) |
| Men | 16 (59.26%) | 21 (63.64%) |
| Body-mass Index, kg/m<sup>2</sup> | 31.44 (7.49) | 28.94 (5.25) |
| **Risk factors** |  |  |
| Smoker | 2 (7.41%) | 5 (15.62%) |
| Former smoker | 3 (11.11%) | 7 (21.88%) |
| Ethylism | 0 (0.00%) | 4 (12.50%) |
| Chronic obstructive pulmonary disease | 1 (3.70%) | 4 (12.12%) |
| Cardiac insufficiency | 1 (3.70%) | 1 (3.03%) |
| Ischemic cardiopathy | 0 (0.00%) | 3 (9.09%) |
| Occlusive arteriopathy of the lower limbs | 0 (0.00%) | 1 (3.03%) |
| Atrial fibrillation arrhythmia | 1 (3.70%) | 2 (6.06%) |
| Cirrhosis | 0 (0.00%) | 1 (3.03%) |
| Diabetes | 8 (29.63%) | 14 (42.42%) |
| Cancer | 1 (3.70%) | 2 (6.06%) |
| Human immunodeficiency virus | 0 (0.00%) | 1 (3.03%) |
| **Clinical history** |  |  |
| Origin |  |  |
| Home | 3 (11.11%) | 4 (12.12%) |
| Surgery Ward | 7 (25.93%) | 8 (24.24%) |
| Hospital Ward | 12 (44.44%) | 11 (33.33%) |
| Other Hospital | 5 (18.52%) | 10 (30.30%) |
| Length of stay intensive care unit | 12.00 (8.50-23.00) | 17.00 (12.00-35.00) |
| Length of stay hospitalisation | 26.00 (16.50-37.50) | 26.00 (19.00-38.25) |
| Discharge modalities |  |  |
| Home | 12 (44.44%) | 11 (33.33%) |
| Rehabilitation facility | 15 (55.56%) | 22 (66.67%) |
| **Clinical parameters** |  |  |
| IGS II | 37.77 (12.19) | 43.13 (15.39) |
| Mechanical ventilation duration | 18.00 (7.00-25.50) | 18.00 (9.25-30.25) |
| PaO<sub>2</sub>/FiO<sub>2</sub> | 121.00 (91.00-153.00) | 97.00 (80.00-165.00) |

Mean MDP at three months was 26.15 (SD 15.48) in the ETR group vs. 44.76 (SD 19.25) in the SP group. Mean difference between groups was -18.61 (95% CI -27.78 to -9.44; p<0.0001) and was statistically significant. MDP total score distribution between groups is presented in figure 2.

**Figure 2.**
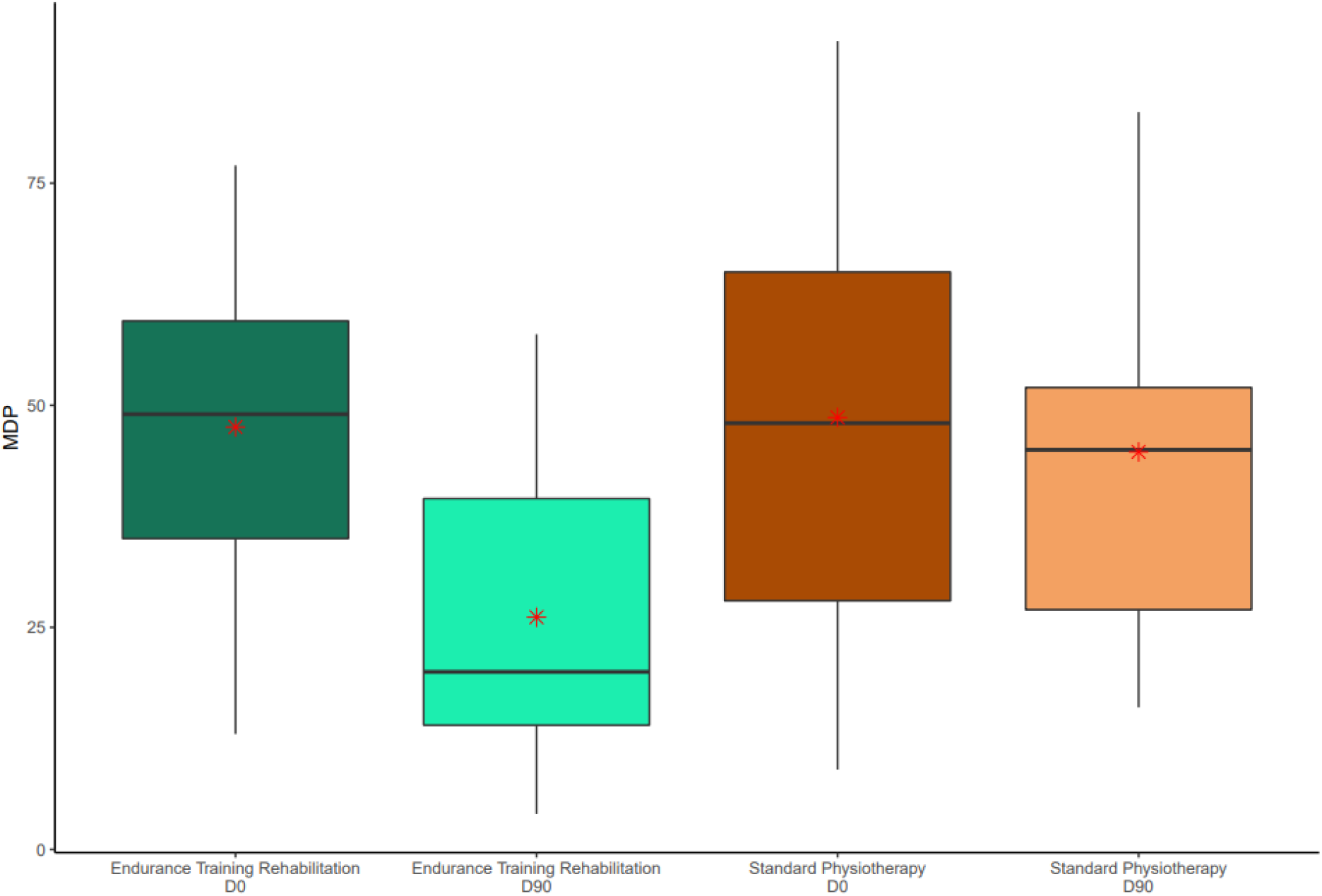
Total MDP comparison between groups at baseline and day-90. The middle horizontal line represents the median; outliers are displayed as separate black points. The outside values are smaller than the lower quartile minus 1·5 times the interquartile range or are larger than the upper quartile plus 1·5 times the interquartile range. Means are represented by a red cross. Abbreviations: MDP: Multidimensional Dyspnea Profile; ETR: endurance training rehabilitation; SP: standard physiotherapy; D0: Day-0; D90: Day-90

Mean breathing discomfort, sensory dimension and emotional response at three months were 3.44 (SD 2.10), 10.59 (SD 8.90) and 12.11 (SD 9.88) respectively in the ETR group versus 5.18 (SD 2.02), 20.52 (SD 9.33) and 19.06 (11.82) respectively in the SP group. In these MDP dimensions a statistical significant difference was found at three months and the mean difference were -1.74 (95% CI -2.81 to -0.67; p=0.0006), -9.92 (95% CI -14.67 to -5.18; p<0.0001) and -6.95 (95% CI -12.66 to -1.24; p=0.0140) respectively for breathing discomfort, sensory dimension and emotional response. Distribution of these dimensions are presented in figure 3. Full data about MDP primary outcome are available in table 2.

**Table 2:** Main outcomes. Data are mean (SD). Differences are expressed as mean (IC95%) Abbreviations: MDP: Multidimensional Dyspnea Profile; ETR: endurance training rehabilitation; SP: standard physiotherapy; D0: Day-0; D90: Day-90

|  | Baseline |  |  |  | D-90 |  |  |  |
| --- | --- | --- | --- | --- | --- | --- | --- | --- |
|  | ETR | SP | Difference | p value | ETR | SP | Difference | p value |
| **MDP** |  |  |  |  |  |  |  |  |
| **Total score** | 47.56 (16.10) | 48.64 (24.35) | -1.08<br>(-12.13 to 9.96) | 0.85 | 26.15 (15.48) | 44.76 (19.25) | -18.61<br>(-27.78 to -9.44) | <0.0001 |
| **Breathing discomfort** | 5.33 (1.94) | 5.21 (1.85) | 0.12<br>(-0.86 to 1.10) | 0.81 | 3.44 (2.10) | 5.18 (2.02) | -1.74<br>(-2.81 to -0.67) | 0.0006 |
| **Sensory dimension** | 18.96 (7.66) | 20.45 (12.64) | -1.49<br>(-6.80 to 3.82) | 0.85 | 10.59 (8.90) | 20.52 (9.33) | -9.92<br>(-14.67 to -5.18) | <0.0001 |
| **Emotional response** | 22.89 (11.67) | 22.97 (13.17) | -0.08<br>(-6.59 to 6.42) | 0.98 | 12.11 (9.88) | 19.06 (11.82) | -6.95<br>(-12.66 to -1.24) | 0.0140 |
| **mMRC** | 2.37 (0.63) | 2.30 (0.59) | 0.07<br>(-0.25 to 0.38) | 0.65 | 1.33 (0.62) | 2.09 (1.10) | -0.76<br>(-1.21 to -0.30) | 0.001 |
| **SF-12** |  |  |  |  |  |  |  |  |
| **Total score** | 74.52 (13.10) | 70.47 (11.21) | 4.05<br>(-2.29 to 10.39) | 0.21 | 83.36 (14.97) | 75.13 (15.79) | 8.24<br>(0.22 to 16.25) | 0.14 |
| **Physical dimension** | 35.10 (6.55) | 31.45 (5.98) | 3.64<br>(0.37 to 6.92) | 0.0297 | 39.76 (8.57) | 32.80 (8.15) | 6.95<br>(2.62 to 11.29) | 0.016 |
| **Mental dimension** | 40.57 (8.46) | 39.03 (10.80) | 1.54<br>(-3.62 to 6.71) | 0.55 | 45.08 (11.05) | 43.02 (10.88) | 2.06<br>(-3.63 to 7.75) | 0.72 |

**Figure 3.**
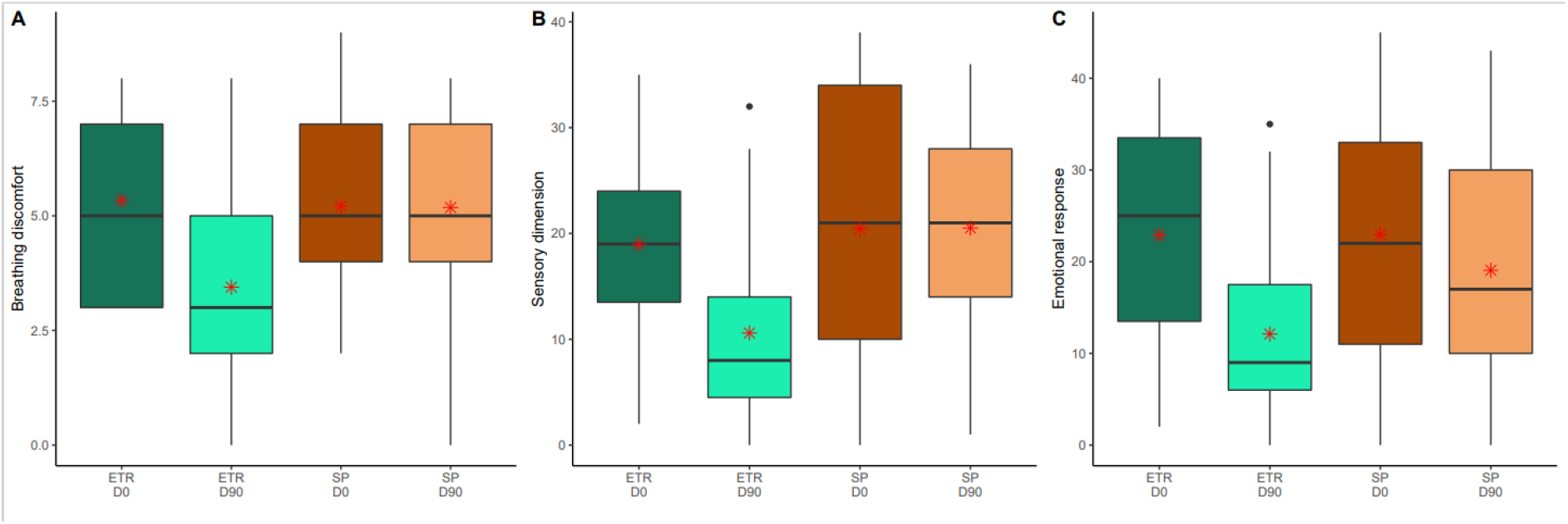
MDP dimensions comparison between groups at baseline and day-90. Panel A refers to breathing discomfort, Panel B refers to sensory dimension and panel C refers to emotional response. The middle horizontal line represents the median; outliers are displayed as separate black points. The outside values are smaller than the lower quartile minus 1·5 times the interquartile range or are larger than the upper quartile plus 1·5 times the interquartile range. Means are represented by a red cross. Abbreviations: MDP: Multidimensional Dyspnea Profile; ETR: endurance training rehabilitation; SP: standard physiotherapy; D0: Day-0; D90: Day-90

Mean mMRC at three months was 1.33 (SD 0.62) in the ETR group and 2.09 (SD 1.10) in the SP group, a statistically significant difference (mean difference -0.76 [95% CI -1.21 to -0.30; p=0.001]).

Mean SF-12 total scores at the end of the intervention period were comparable between ETR and SP groups (mean difference 8.24 [95% CI 0.22 to 16.25]; p=0.14).

Separate analysis of the physical dimension of SF-12 score retrieved a significant difference between groups (mean difference 6.95 [95% CI: 2.62 to 11.29]; p=0.016). No statistically significant difference was observed between groups in the evolution of the mental dimension of SF-12 score (mean difference 2.06 [95% CI -3.63 to 7.75]; p=0.72). Full details about secondary outcomes are available in table 2.

## Discussion

In this multicentre randomised trial, a three-month ETR course decreased dyspnoea for patients with “long-covid” following CARDS compared to usual care involving standard physiotherapy. A concomitant improvement of the physical component of the SF-12 quality of life scale was observed, without significant modification neither of mental component nor of the global quality of life.

Whereas physiotherapy has been used for a long time, little data exists concerning the procedure’s implementation, leading to an important variability from one physiotherapist to another for a given patient [25–27]. Management of patients with chronic respiratory insufficiency and dyspnoea in ETR is however based on the standardisation of the procedure, allowing accurate evaluation of the treatment effectiveness on breathlessness and quality of life.

Dyspnoea is a broad symptom persisting beyond the acute phase of COVID-19, particularly in women, the elderly and multi-comorbid patients and may worsen in the first post COVID-19 year [28–30]. In our population, only 200 of the 871 (23%) screened patients were little or non-affected by breathlessness during daily activities. Dyspnoea appears to be one of the major symptoms reported in “long covid” [14, 22, 25, 31–33]. Shortness of breath, breathlessness and associated sensation of breathing impairment are described by 40 to 50 % of patients, and more marked in the most severe patients [34]. Such late symptoms are coherent with those usually noted lately during classical ARDS evolution. In patients cured from COVID-19, dyspnoea is, interestingly, opposite to the “happy hypoxemia” which disconnects the severity of hypoxemia and the only mild respiratory discomfort reported by the patients [5]. Reversion of the pattern, associating concomitant improvement of haematosis with worsening of dyspnoea could be explained by compromise of the alveolocapillary membrane, as illustrated by altered DLCO, pulmonary function tests and CT abnormalities reported in a growing number of studies [15, 33, 34]. Moreover, dyspnoea, decreased DLCO and pulmonary fibrosis seem to be closely associated [34]. These functional data are consistent with early histological studies emphasising the importance of local inflammation and diffuse alveolar damages and with known histological evolution of ARDS.

The currently observed improvement of dyspnoea associated with an ETR course is in accordance with previous cohorts analysis, following COPD, moderate severity COVID-19 and both classical ARDS and CARDS [25, 26, 35, 36]. The improvement of physical abilities observed in cohort studies is coherent with the more than 50% decrease of MDP score after a three-month ETR course we have identified. Improvement of dyspnoea was accompanied by a significant improvement of the mMRC, possibly reflecting the growth of functional ability after ETR. This raises the question of the interdependency of dyspnoea and disability. Whether the latter was improved through improved dyspnoea or whether the decrease in perceived dyspnoea was merely a consequence of the growth of functional ability has long been debated and remains a matter for future research [37].

Most studies about pulmonary rehabilitation assess dyspnoea through one-dimensional assessments (Borg scale, mMRC) whereas dyspnoea mechanisms are naturally intricate and multifactorial, and should be assessed on both emotional and sensorial dimensions [38]. We indeed demonstrated a reduction in breathing discomfort, sensory dimension and emotional response to dyspnoea. Moreover, during “long covid”, light is nowadays being shed on dyspnoea’s direct effects on anxiety, depression and quality of life [14, 22, 25, 31, 32, 36]. A beneficial effect of early mobilisation during the ICU stay, is associated with an improvement in functional capacity and may contribute to improving quality of life [39]. Conversely, the benefit from rehabilitation on quality of life beyond initial hospitalisation remains poorly studied [20, 25, 36]. In our work, the mitigation of dyspnoea is associated with an improvement of the physical part of quality of life measured by SF-12 score. The global SF-12 score change did not differ between groups. This may be partly explained by the natural evolution of quality of life over time [25]. Another explanation could be a lack of power. Using a more sensitive evaluation tool, such as the Short Form-36, might have been able to detect a difference, as previously demonstrated in COPD or in COVID-19 [25–27]. The SF-36 was actually our first choice for assessing QOL but, for practical and temporal reasons during the pandemic, we ultimately opted for the more time-efficient SF-12. The relative part of dyspnoea within multiple chronic symptoms is difficult to establish. A large combination of organ dysfunctions, not affected by physiotherapy, such as ageusia, headaches, hair loss, intestinal disorders may play a major part in global quality of life, especially in a young population with few comorbidities [14]. Another explanation suggested by Grosbois et al. would be that anxiety and depression (mental component of SF-12) seem to have a greater impact on dyspnoea compared than the physical component [27]. Therefore, an improvement in dyspnoea could have a greater effect on the physical rather than on the mental part of QOL.

To the best of our knowledge, this is the first randomised controlled work studying the effect of endurance training rehabilitation in CARDS patients, which is a major strength of our study. Furthermore, we chose to collect the outcomes by an independent assessor, blinded to the intervention, in order to minimise the biases due to the open nature of this type of trial. As standard physiotherapy is equally recommended, we designed the protocol to allow the non-intervention group to benefit from physiotherapy and still offered the opportunity of ETR after the 3 months evaluation, considering the recent guidelines [24]. Finally, the formalisation of intervention, using existing guidelines, and the recruitment of rehabilitation expert physiotherapists allowed us to expect a reproducible training, reinforcing the reproducibility of the method in any location [23, 24].

Our study has some limitations. First, the impossibility of reaching the 200 planned participants forced us to prematurely interrupt the study. However, our sample size estimation was based on a rather pessimistic expected MDP difference of 12 with a large SD of 30. The observed difference was actually larger with a narrower SD. Using our results to calculate the sample size would have given us a number of subjects to include of 40. Second, vaccination and better overall control of the contagion, while fortunately reducing the pandemic, led to a decrease in the possibilities of including patients in the study. Third, we faced a major challenge in referral to pulmonary rehabilitation practices. Indeed, pulmonary rehabilitation practices, usually reserved for COPD, are scarce and no official directory of these rare practices is available. Those latter were eventually saturated, despite the slowing of pandemic, with a surge of both usual COPD patients and “long-covid” patients. It appears that the scarcity of such resources, already contributing to the difficulties of first-line care, also played a role in the availability of post-hospitalisation care [40]. Finally, 53 patients could not be included because they live too far from a rehabilitation practice.

Patients from both groups had an overall good compliance with the rehabilitation procedures and all of them completed their therapy. Second, as mentioned earlier, ETR and physiotherapy, if delivered by the same physiotherapist, differ substantially by the protocolisation of the therapy. Indeed, the administered physiotherapy varies greatly depending on the therapist administering it, whereas ETR is highly protocolised. Hence, the results of ETR are more consistent and homogeneous when compared with SP which currently represents standard of care but failed to improve dyspnoea or QOL in this trial. However, the adequation of physiotherapists practice with guidelines between sessions was not evaluated.

A six-week ETR program did improve dyspnoea and the physical dimension of quality of life amongst patients remaining dyspnoeic after a COVID-19-related acute respiratory distress syndrome, compared to standard physiotherapy. ETR could therefore be the most appropriate treatment for patients presenting dyspnoea after COVID-19, provided rehabilitation practices are available nationwide.

## Data Availability

All data produced in the present study are available upon reasonable request to the authors

## Contributors

This manuscript was initially drafted by CR, JW & FP. GC designed the statistical plan & AF did the statistical analysis. CB, FP, CR, PL, GP & FP contributed to patient selection. CR & PL included patients. All authors contributed to data interpretation, and critical review and revision of the manuscript. All authors had full access to all the data in the study and had final responsibility for the decision to submit for publication.

## Acknowledgments

The authors thank the members of the clinical research centre for their constant help throughout the trial.

